# Genomics-informed Neuropsychiatric Care for Neurodevelopmental Disorders: Results from A Multidisciplinary Clinic

**DOI:** 10.1101/2024.05.08.24307074

**Authors:** Aaron D. Besterman, David J. Adams, Nicole R. Wong, Benjamin N. Schneider, Sunil Mehta, Charlotte DiStefano, Rujuta B. Wilson, Julian A. Martinez-Agosto, Shafali S. Jeste

**Affiliations:** University of California San Diego Department of Psychiatry, Division of Child and Adolescent Psychiatry, San Diego, California, USA; Rady Children’s Hospital of San Diego, San Diego, California, USA; Rady Children’s Institute for Genomic Medicine, San Diego, California, USA, University of California Los Angeles, Division of Child and Adolescent Psychiatry, Department of Psychiatry, Los Angeles, California, USA; University of California Los Angeles, David Geffen School of Medicine, Los Angeles, California, USA; University of California Los Angeles, Semel Institute for Neuroscience and Human Behavior, Los Angeles, California, USA; Department of Psychiatry and Behavioral Sciences, Johns Hopkins School of Medicine, Baltimore, MD, USA., PrairieCare Medical Group, Rochester, Minnesota, USA; Children’s Hospital Los Angeles, Department of Psychiatry, Los Angeles, California, USA; University of Southern California, Department of Psychiatry & The Biobehavioral Sciences, Los Angeles, California, USA; University of California Los Angeles, Department of Human Genetics, Los Angeles, California, USA; Children’s Hospital Los Angeles, Division of Neurology, Los Angeles, California, USA; University of Southern California, Departments of Pediatrics and Neurology, Los Angeles, California, USA

**Keywords:** Neurodevelopmental disorders, Autism Spectrum Disorders, Intellectual Disabilities, Genomic Medicine, Precision Medicine, Genetic Testing, Neuropsychiatric Disorders, Health Services

## Abstract

**Background:** Patients with neurodevelopmental disorders (NDDs) have high rates of neuropsychiatric comorbidities that can be highly impairing and treatment refractory. Genomic medicine may help guide care, as pathogenic variants are identified in up to 50% of patients with NDDs. We evaluate the impact of a genomics-informed, multidisciplinary, neuropsychiatric specialty clinic on the diagnosis and management of patients with NDDs.

**Methods:** We performed a retrospective study of 316 patients from the UCLA Care and Research in Neurogenetics Clinic, a genomics-informed multidisciplinary clinic composed of psychiatry, neurology, medical genetics, psychology, and social work.

**Results:** We observed high rates of psychiatric and medical comorbidity. Among the 246 patients that underwent genetic testing, 41.8% had a pathogenic or likely pathogenic (P/LP) variant. Patients had 62 different genetic diagnoses, with 12 diagnoses shared by two or more patients, including Duplication 15q syndrome (9.18%), Tuberous Sclerosis Complex (3.48%), and Angelman syndrome (1.27%), while 50 diagnoses were found in only single patients. Genetic diagnosis resulted in direct changes to clinical management in all patients with a P/LP variant, including high rates of cascade testing (30.6%), family counseling (22.2%), medication changes (13.9%), clinical trial referral (2.8%), medical surveillance (30.6%), and specialty referrals (69.4%).

**Conclusions:** A genomics-informed model can provide significant clinical benefits to patients with NDDs, directly impacting management across multiple domains for most diagnosed patients. As precision treatments for NDDs advance, establishing a genetic diagnosis will be critical for proper management. With the growing number of rare neurogenetic disorders, clinician training should emphasize core principles of genomic medicine over individual syndromes.

## BACKGROUND

The genetic etiology of neurodevelopmental disorders (NDDs), such as autism spectrum disorders (ASD), intellectual disability (ID), and global developmental delay (GDD), includes a combination of common, low effect-size variants and rare, highly-pathogenic variants that disproportionately impact early brain development [1,2]. Genetic testing with next-generation sequencing, such as exome or genome sequencing, allows for the detection of rare pathogenic variants in up to 50% of severely affected patients with NDDs [3] and is now recommended as the first line diagnostic test for patients with congenital anomalies or ID [4]. A genetic diagnosis can lead to improved medical monitoring, more accurate reproductive counseling, increased social and psychological support through patient advocacy groups, refined prediction of natural history and prognosis, and access to disorder-specific research [5]. However, it remains unclear which clinical care delivery model for patients with NDDs is most compatible with a genomics-informed approach.

Traditionally, clinical management has fallen on primary care physicians, with consultation from a patchwork of subspecialists, with many patients and families struggling to find providers willing or able to care of patients with NDDs and their complex needs. The “patient-centered medical home” model of care, defined as “…a model for strengthening primary care through the reorganization of existing practices to provide patient-centered, comprehensive, coordinated, and accessible care that is continuously improved through a systems-based approach to quality and safety” [6], alleviates some of the limitations of the traditional medical model by streamlining services and centralizing subspecialist recommendations. Another approach to clinical care for patients with NDDs are disease-specific specialty clinics at tertiary care medical centers that focus on relatively common NDDs like Down syndrome [7] or Tuberous Sclerosis Complex [8]. These clinics, often supported by patient advocacy groups, provide patients and families with a genetics-first approach to care and access to highly trained specialists for a specific disorder or a group of related disorders. However, this model still leaves the majority of patients with NDDs with an ongoing and daunting “diagnostic and therapeutic odyssey” for both medical and neuropsychiatric care [9–11]. Patients from our clinic have reported particular difficulty with delays between initial caregiver concerns for NDDs and formal developmental or genetic diagnoses and barriers to specialized care including long wait times for an appointment, lack of insurance coverage, lack of availability of local evaluations, transportation difficulties, and native language differences [12].

General pediatric neurology and psychiatry clinics may offer neuropsychiatric care for these patients, but traditional clinic models are symptom-focused and lack universal genetic testing or a genomics-informed approach to diagnosis, prognosis, and treatment. Given the enormous burden of neuropsychiatric comorbidities for patients with NDDs [13] and the growing number of NDDs with an identifiable genetic etiology [1,2], genome-informed neuropsychiatric care for patients with a known or suspected genetic condition is warranted.

We describe a novel delivery model of neuropsychiatric care for patients with known or suspected neurogenetic conditions at the University of California, Los Angeles (UCLA), the UCLA Care and Research in Neurogenetics (CARING) Clinic. The clinic provided specialty services in medical genetics, neurology, psychiatry, and psychology. We describe our experience over a five-year period, evaluating 316 patients. We identify key ways in which our genomics-informed approach to neuropsychiatric care can alleviate aspects of the diagnostic and treatment odyssey [12], and highlight challenges that we faced. Based on our experience, we highlight the urgent need for medical educators and health care administrators to reconsider disorder-specific approaches to genomic medicine and instead emphasize more generalized genomic training so that clinicians and health systems are equipped to expertly care for a growing population of patients with ultra-rare genetic disorders.

## METHODS

Research approval was granted by the UCLA Medical Institutional Review Board 3. One-hundred-ten patients and/or their legal guardians provided informed consent for prospective collection of clinical data (UCLA Institutional Review Board (IRB) #: 14-001908). Patient assent was obtained when feasible. With an IRB-approved waiver of consent, the charts of an additional 206 patients were retrospectively reviewed (UCLA IRB#: 19-000121). Upon manual extraction from the UCLA electronic medical record (D.J.A., N.R.W.), all patient data was coded into an encrypted database. The code key with personally identifiable information was stored separately from the database in a secure, HIPAA-compliant cloud platform. Only members of the research team responsible for data extraction had access to the code key. All subsequent analyses were conducted using the coded, de-identified data. Statistical analyses were performed using SPSS version 29.0.1.1.

## RESULTS

### Demographics and Family Structure

Subjects were evaluated in the UCLA CARING Clinic between January 1, 2014, and January 1, 2019. The cohort consisted of 316 patients, with 65.8% assigned male at birth (Fig. S1A). The patients had a mean age at intake of 118.99 months (*SD* = 92.7, Fig. S1B). One-hundred-seventy-eight patients (56.3%) self-identified as white, 55 (17.4%) Hispanic or Latino, 42 (13.3%) as Asian, six (1.9%) as Black or African American, and two (0.6%) as American Indian or Alaska Native. Thirty-three (10.44%) identified as another, multiple, or unknown ethnic background (Fig. S1C). Based on address of residence, we determined Area Deprivation Index [14] (ADI) and Social Vulnerability Index [15] (SVI) as measures of socioeconomic status (SES). The ADI is reported as a national percentile of socioeconomic disadvantage, with lower values indicating higher SES; the SVI is reported as a value between zero and one, with lower values again indicating higher SES. The cohort had an average ADI national percentile of 9.6 (*SD* = 11.9) and an average SVI of 0.35 (*SD* = 0.2), relatively low levels of disadvantage (Fig. S2). We compared the mean ADI and SVI between patients who received and did not receive exome or genome sequencing using the Mann-Whitney U Test due to non-normality, and did not find any significant difference (p = 0.11 and 0.34, respectively), suggesting similar access to these tests across SES. Similarly, we did not see any correlation between ADI or SVI and the number of psychiatric diagnoses per patient (r = −0.02, p = 0.75; r = −0.04. p = 0.48, respectively).

Fifty-six patients (17.7% of the total cohort) had a sibling with an NDD or other psychiatric diagnosis. Twenty-six of the 56 (8.2% of the total cohort) had a sibling with an NDD only. Fifteen of the 56 (4.7% of the total cohort) had a sibling with just a psychiatric diagnosis, and 15 of the fifty-six (4.7% of the total cohort) had a sibling with both an NDD and psychiatric diagnosis. Four of the 56 families with multiple affected children had more than one child evaluated in the CARING clinic.

### Clinical Assessment and Management

Patients were referred to the UCLA CARING Clinic from many settings, including physicians within the UCLA Health system, outside physicians, patient advocacy groups (*e.g.,* Dup15Q Alliance), and research studies. Patients encountered by CARING Clinic providers in other settings (i.e., research laboratories or other UCLA clinics) were often brought to CARING for further multidisciplinary assessment and treatment by other CARING clinicians. Some families were self-referred, with an increasing number of these referrals as the clinic became known in the community.

Upon intake, patients were evaluated over one or several monthly visits by specialists in neurology, medical genetics, psychiatry, and/or clinical psychology (Fig. 1). Although formal neuropsychological testing was not routinely completed, great care was taken to capture a detailed developmental and clinical history to refine and update neurodevelopmental, psychiatric, and neurological diagnoses. On admission, most patients had a diagnosis of ASD, ID, or both (Fig. 2A), and a significant number of new diagnoses were made by CARING Clinic clinicians (Figs. 2B, 2C). Most patients also had a significant history of development delays, mostly commonly combined motor and language delays, at time of referral (Fig. 2D). Both psychiatric (*e.g.,* anxiety, depression) (Table 1) and neurologic disorders (*e.g.,* epilepsy, spells, and cerebral palsy) (Fig. 3) were common in clinic patients. All genetics evaluations, including dysmorphology exams, pedigrees, and informed consent for genetic testing were completed by a single medical geneticist (J.A.M.A.) specializing in NDDs without the assistance of a genetic counselor.

**Fig. 1.**
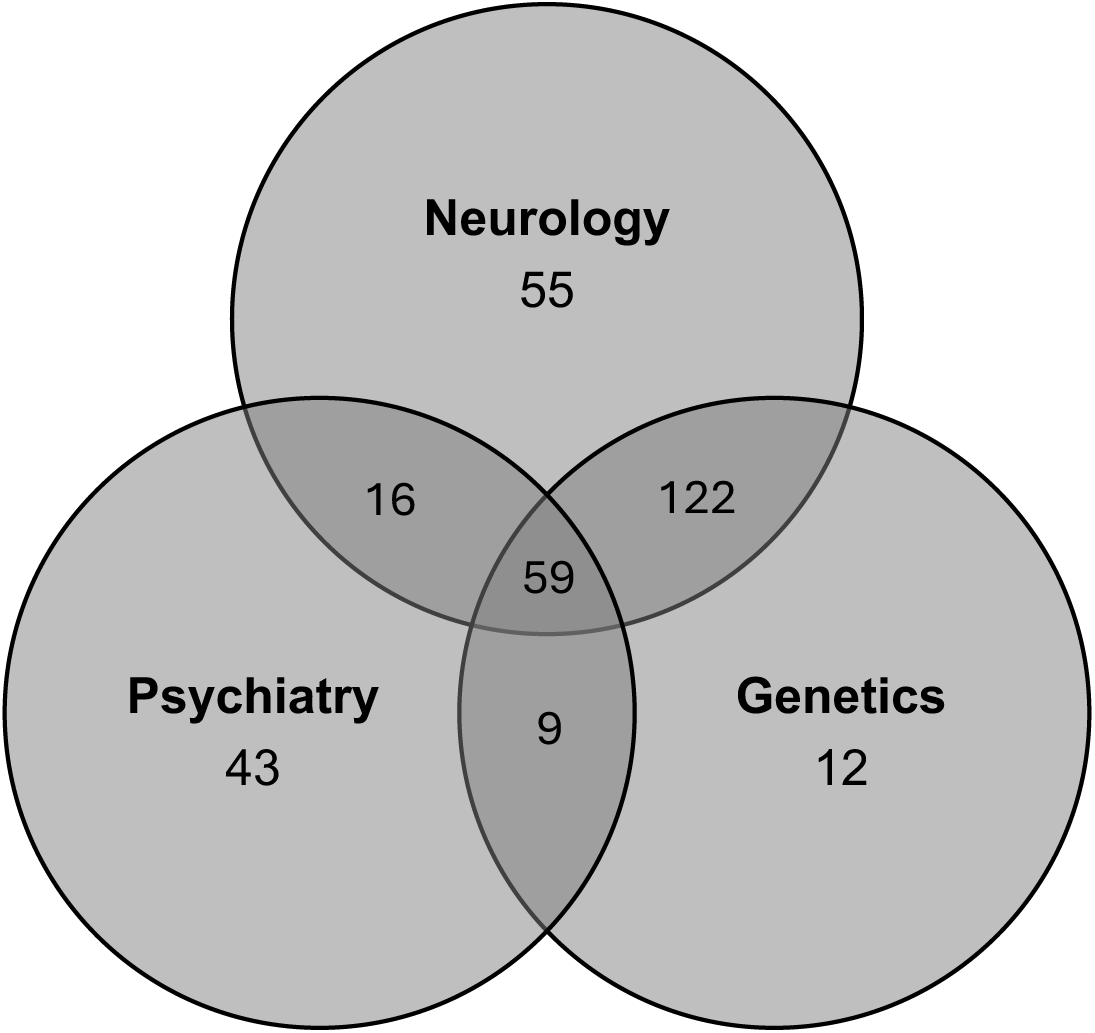
Number of Patients Seen by Each CARING Clinic Physician Specialty. Sum of all numbers within each circle provides the total number of patients seen by each physician specialty. The numbers within regions of overlap represent the number of patients seen by each combination of physician specialists. All patients seen by social work (*n* = 61) and clinical psychology (*n* = 44) were seen by at least one physician in the clinic.

**Fig. 2.**
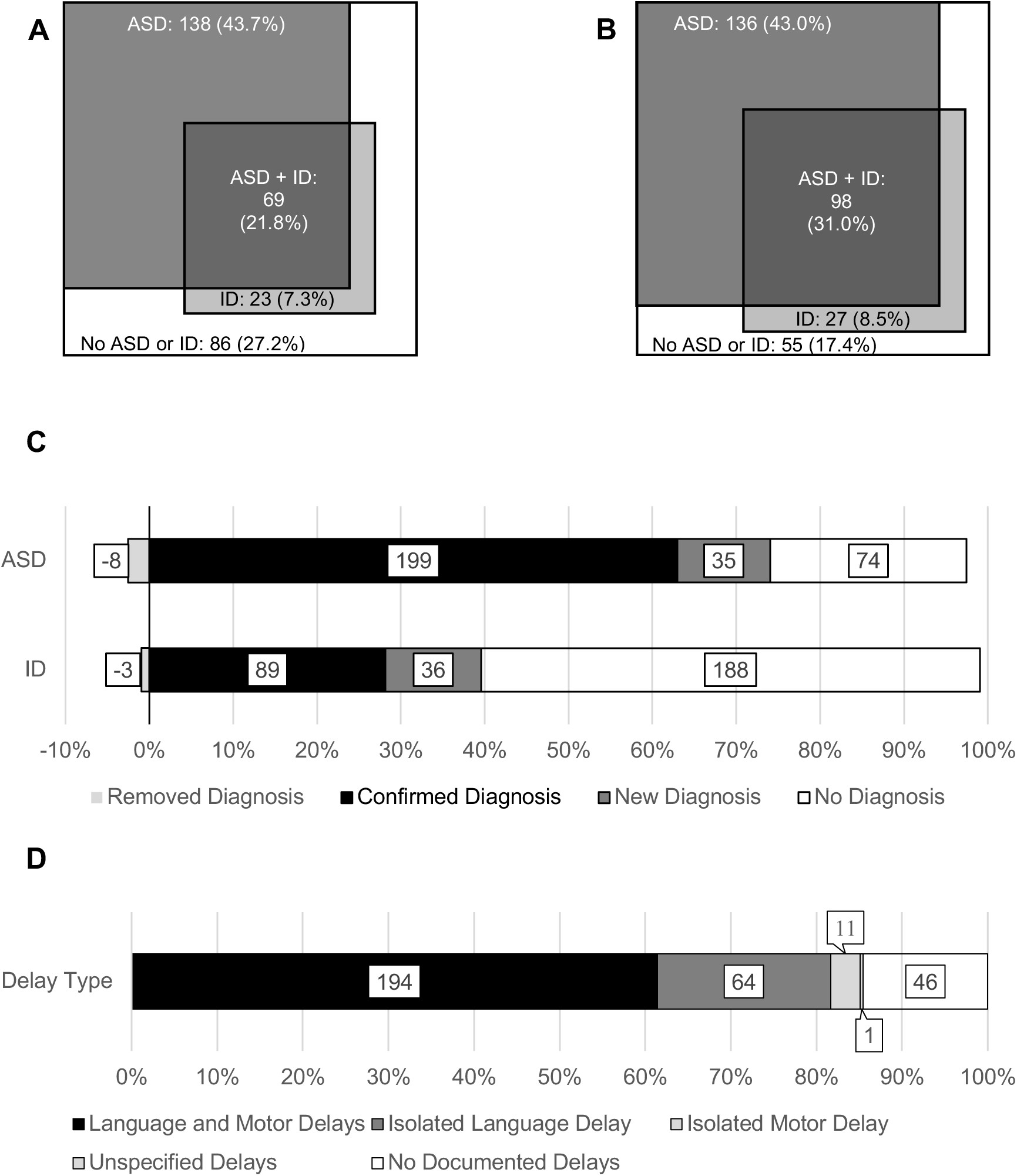
Neurodevelopmental Disorder Diagnoses Before and After Clinic Intake. **A** Nested diagram of ASD and ID diagnoses prior to clinic intake, **B** Nested diagram of ASD and ID diagnoses after clinic intake. For both A and B, the outer squares represent all patients within the cohort. **C** ASD and ID diagnoses removed, confirmed, or added by the CARING Clinic. **D** Types of developmental delays ever experienced by CARING Clinic patients. “Diagnosed Prior to Intake” is defined as a diagnosis prior to the patient’s first appointment in the CARING Clinic. “Diagnosed after Intake” is defined as a diagnosis after the patient’s first appointment in the CARING Clinic. The sum of “Diagnosed Prior to Intake” and “Diagnosed After Intake” represents the number of patients ever being diagnosed with each neurodevelopmental disorder. Disorders are listed in descending order by frequency. *Abbreviations:* ASD, autism spectrum disorders; GDD, Global Developmental Delay; ID, Intellectual Disability

**Fig. 3.**
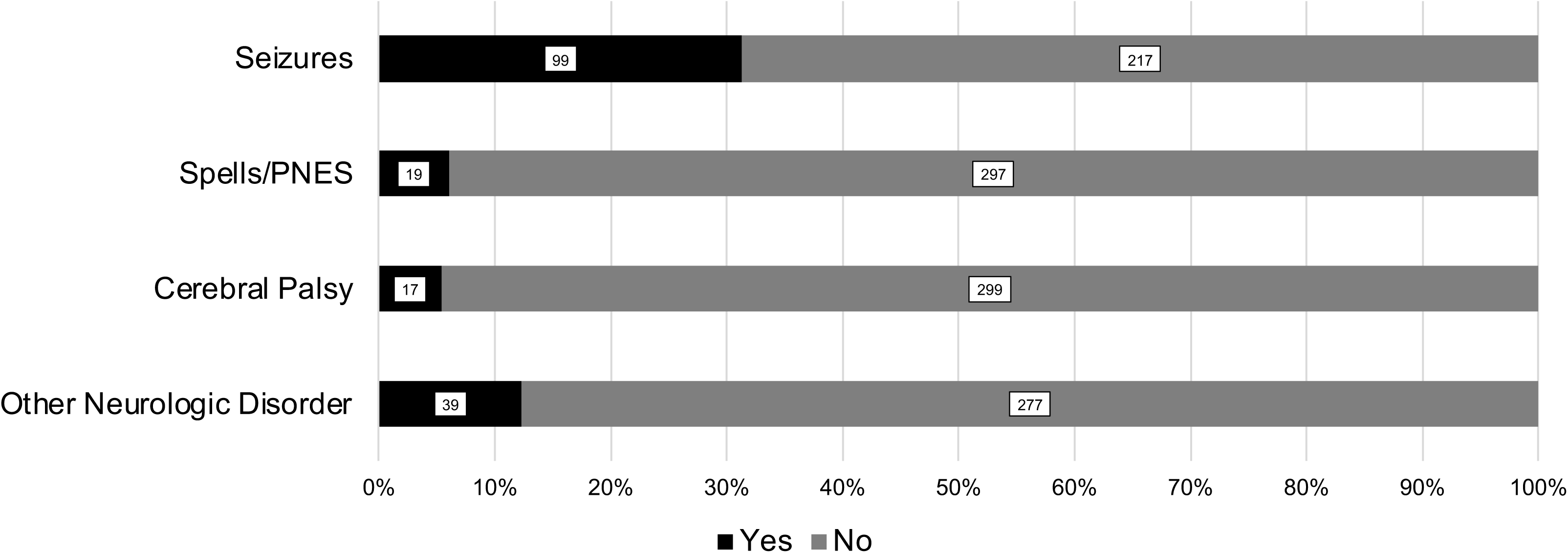
Neurologic Disorder Diagnoses. Number within bar represents absolute number of patients with specified diagnosis. *Abbreviations:* PNES, psychogenic nonepileptic seizures

**Table 1.** Psychiatric Disorders Diagnosed Before and After Clinic Intake.

| Psychiatric Disorder | Prior to Intake | After Intake | Total |
| --- | --- | --- | --- |
| Attention-Deficit/Hyperactivity Disorder | 68 (21.52%) | 48 (15.19%) | 116 (36.71%) |
| Anxiety Disorder | 55 (17.41%) | 41 (12.97%) | 96 (30.38%) |
| Depressive Disorder | 19 (6.01%) | 9 (2.85%) | 28 (8.86%) |
| Obsessive-Compulsive or Related Disorder | 18 (5.70%) | 5 (1.58%) | 23 (7.28%) |
| Tic Disorder | 13 (4.11%) | 1 (0.32%) | 14 (4.43%) |
| Disruptive, Impulse-Control, or Conduct Disorder | 6 (1.90%) | 7 (2.22%) | 13 (4.11%) |
| Schizophrenia Spectrum/Psychotic Disorder | 7 (2.22%) | 1 (0.32%) | 8 (2.53%) |
| Bipolar Disorder | 7 (2.22%) | 0 (0.00%) | 7 (2.22%) |
| Substance Use Disorder | 1 (0.32%) | 1 (0.32%) | 2 (0.63%) |
| Anorexia Nervosa | 1 (0.32%) | 0 (0.00%) | 1 (0.32%) |
| Gender Dysphoria | 1 (0.32%) | 0 (0.00%) | 1 (0.32%) |

CARING patients had a high burden of psychoactive medications, with an average of 1.2 psychoactive medication classes (*SD* = 1.5; Fig. S3) on intake compared to 0.69 non-psychoactive medications (*SD* = 1.42, range 0-8) and 0.5 dietary supplements (*SD* = 1.3, range 0-9) (Table S2). CARING Clinic clinicians recommended medication changes (*e.g.,* initiation, discontinuation, or dosage change) for 61.4% of patients on psychotropics. Gastrointestinal and cardiac issues were the most common medical comorbidities (Fig. S4). Referrals were frequently made by CARING clinicians to other neurodevelopmental (Table S2) and medical (Table S3) specialists. Nearly 70% of patients had received occupational therapy and speech therapy and nearly 50% had received physical therapy and applied behavioral analysis prior to clinic intake (Fig. S5). An additional 5-10% of patients received referrals for these services after CARING clinic evaluations. Clinical psychology consultations focused on evaluating the patient’s educational and therapeutic activities and making recommendations for appropriate additions or modifications. Communication skills were evaluated and recommendations for assistive and alternative communication strategies were provided as appropriate.

### Genetic Testing and Diagnosis

Among all patients in the cohort, 77.9% of patients received genetic testing before, during, and/or after CARING Clinic (Fig. 4). Most patients who did not receive genetic testing were either lost to follow-up or had the request denied by insurance due to the patients not meeting individual insurance policy requirements for testing, which can vary widely and be inconsistently applied in the United States. Chromosomal microarray testing was the most common test that patients received, followed by exome sequencing and then fragile X testing (Fig. 5). The largest percent increase in testing following a visit to CARING clinic was for whole exome sequencing (Fig. 5), driven by the availability of institutional clinical testing since 2012. Based on professional guidelines [16], genetic variants were classified as pathogenic, likely pathogenic, likely benign, or of uncertain clinical significance. A pathogenic or likely pathogenic (P/LP) variant was considered a “positive” result, a likely benign variant, or a variant of uncertain significance, was considered an “inconclusive” result, and no reported variant was considered a “negative” result. Among all tested, 41.8% of patients had a positive result,13.0% had an inconclusive result, and 16.8% had a negative result genetic testing (Fig. 4). CARING Clinic patients had 62 different genetic diagnoses, with twelve genetic diagnoses being shared by two or more patients (Table 2) and 50 genetic diagnoses being carried by only a single patient (Table S4).

**Fig. 4.**
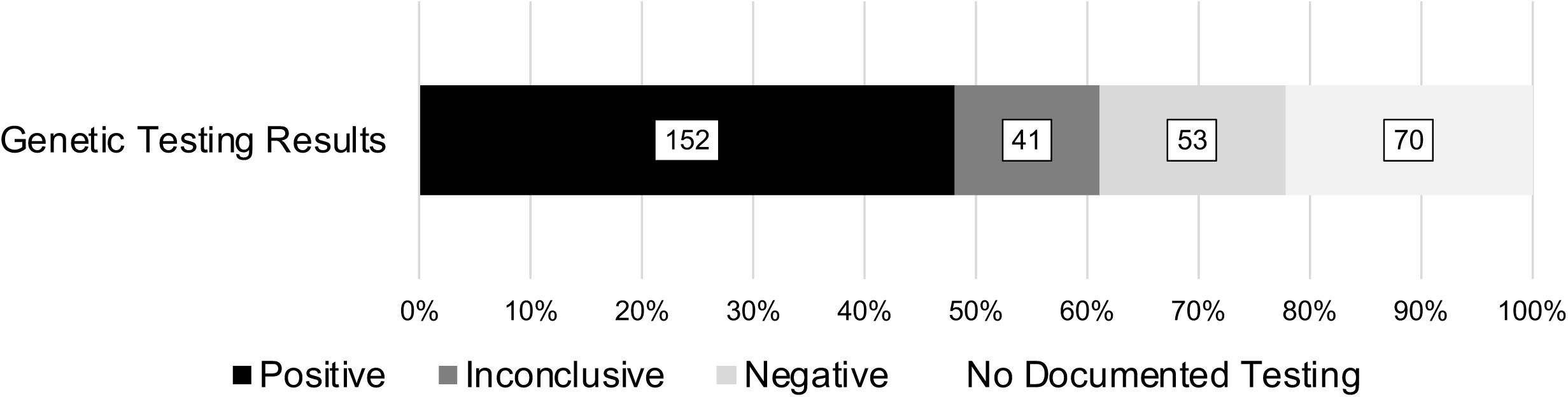
Genetic Test Results for Patients Evaluated in CARING Clinic. Number within bar represents absolute number of patients with specified genetic testing outcome or status. Values include testing that occurred both before and after clinic evaluations.

**Fig. 5.**
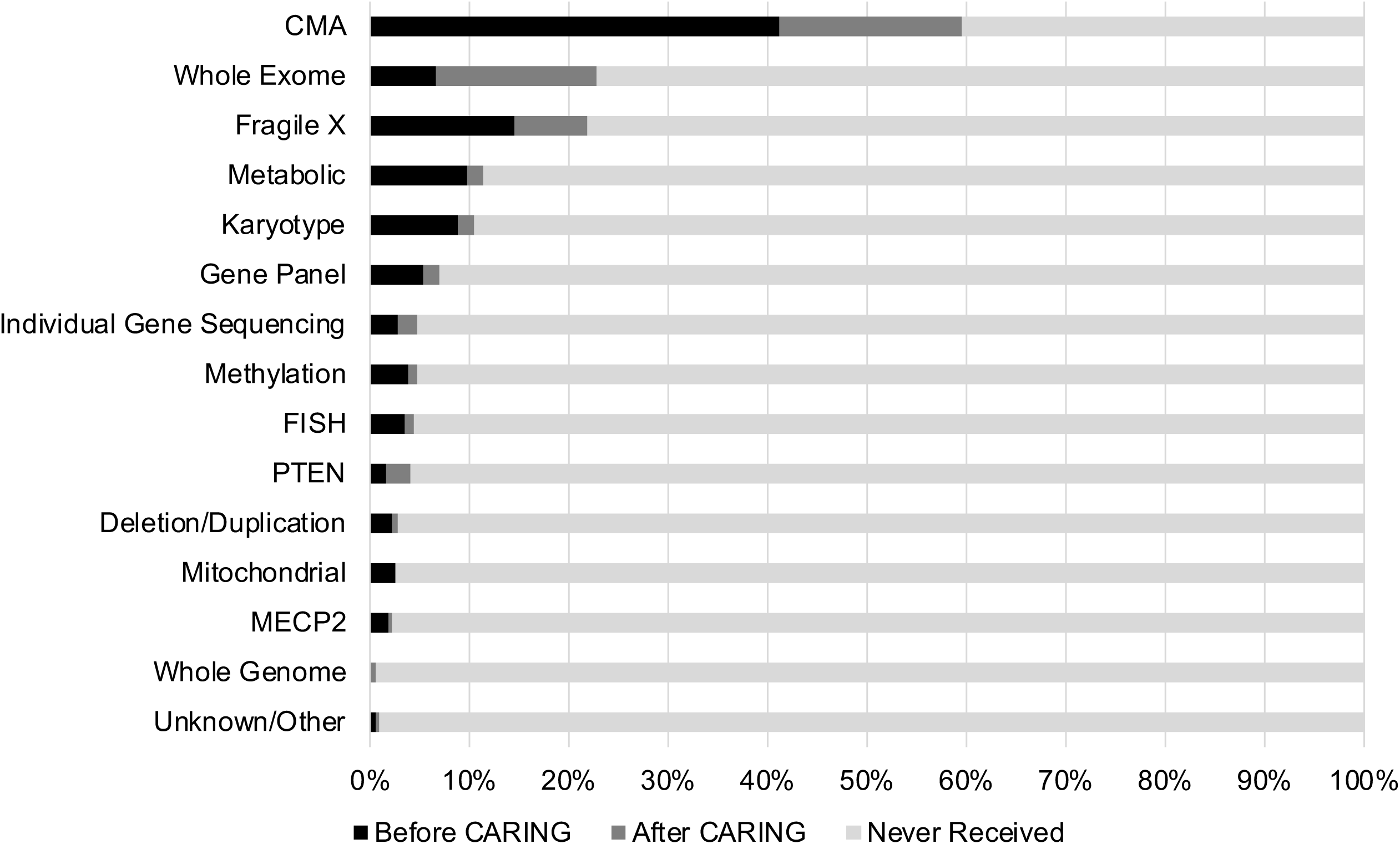
Genetic Tests Before and After CARING Clinic. Bars represent the percent of patients who received each type of genetic tests before and after being evaluated at the CARING Clinic. *Abbreviations*: CMA, chromosomal microarray; FISH, Fluorescence in situ hybridization; PTEN, phosphatase and tensin homolog; MECP2, methyl CpG binding protein 2

**Table 2.** Genetic Disorders Shared by Two or More Patients.

| **Genetic Disorder** | **N** | **%** |
| --- | --- | --- |
| Duplication 15q Syndrome* | 29 | 9.18 |
| Tuberous Sclerosis Complex* | 11 | 3.48 |
| Angelman Syndrome* | 4 | 1.27 |
| 15q11.2 BP1-BP2 microdeletion Syndrome* | 3 | 0.95 |
| 22q11.2 Deletion Syndrome* | 3 | 0.95 |
| Multiple Sulfatase Deficiency<sup>#</sup> | 3 | 0.95 |
| Coffin-Siris Syndrome | 2 | 0.63 |
| Fragile X Syndrome | 2 | 0.63 |
| Kabuki Syndrome | 2 | 0.63 |
| Megalencephaly-Capillary Malformation Syndrome | 2 | 0.63 |
| Pitt-Hopkins Syndrome | 2 | 0.63 |
| Prader-Willi Syndrome | 2 | 0.63 |

\*Specialty clinics for these disorders are present at UCLA, # all members within a single family

### Impact on Clinical Management

Thirty-six out of the 152 patients (23.7%) who had a P/LP variant in our clinic received their genetic diagnosis from our clinic, while the rest were referred to us for management after receiving a genetic diagnosis elsewhere. The genetic diagnosis impacted clinical management for all 36 patients with P/LP variants in at least one domain (Tables 3 and S5). New medical specialist referrals were the most common change in management, but there were also high rates of subsequent cascade genetic testing and ongoing medical surveillance for known disorder-associated comorbidities (e.g. seizures, cancers) (Tables 3 and S5). For a small number of patients, there were direct changes in medication management and new enrollments in disorder-based clinical trials (Tables 3 and S5). These changes were based on reported medication effectiveness or potential safety concerns for a specific genetic diagnosis (often from case reports or series). These changes were not due to pharmacogenetic testing results, as this was not a standard part of our clinical practice, given unproven clinical benefits for psychotropic management, especially in youth [17].

**Table 3.**
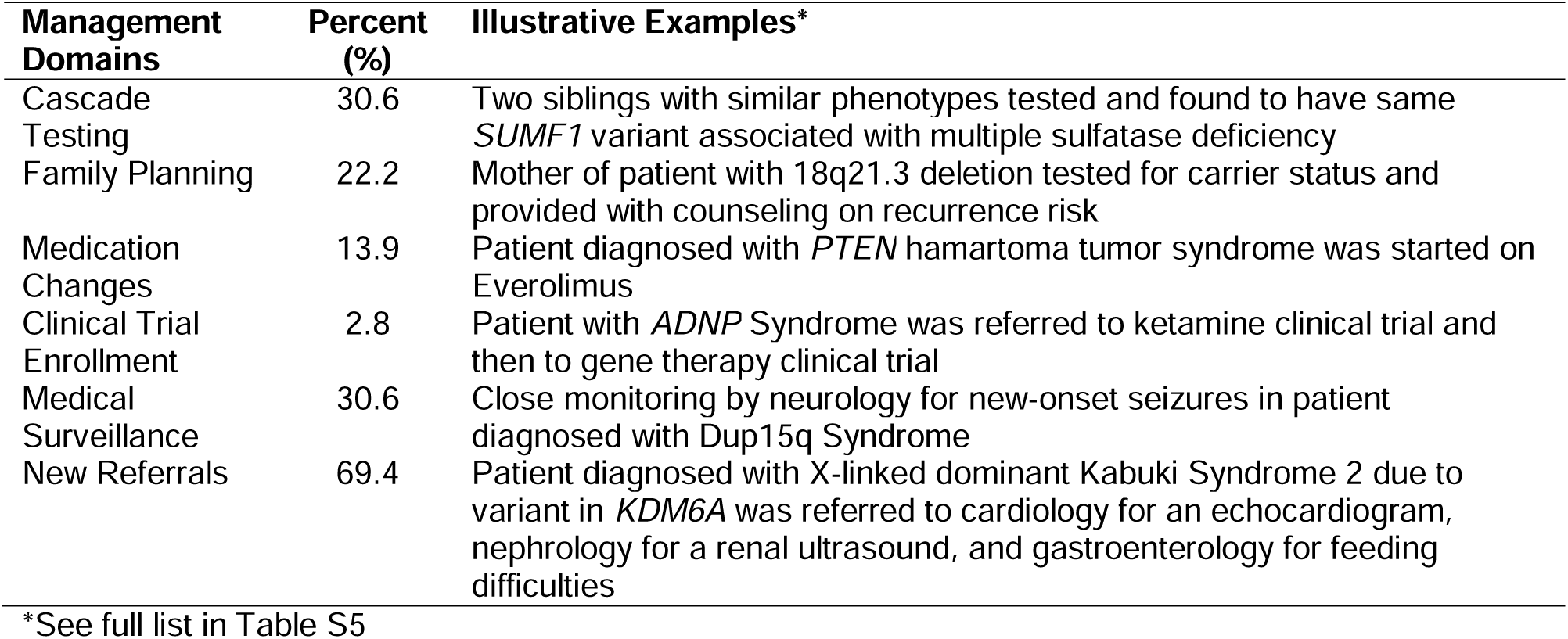
Impact on Clinical Management for Patients with Positive Genetic Findings.

| Management Domains | Percent (%) | Illustrative Examples* |
| --- | --- | --- |
| Cascade Testing | 30.6 | Two siblings with similar phenotypes tested and found to have same *SUMF1* variant associated with multiple sulfatase deficiency |
| Family Planning | 22.2 | Mother of patient with 18q21.3 deletion tested for carrier status and provided with counseling on recurrence risk |
| Medication Changes | 13.9 | Patient diagnosed with *PTEN* hamartoma tumor syndrome was started on Everolimus |
| Clinical Trial Enrollment | 2.8 | Patient with *ADNP* Syndrome was referred to ketamine clinical trial and then to gene therapy clinical trial |
| Medical Surveillance | 30.6 | Close monitoring by neurology for new-onset seizures in patient diagnosed with Dup15q Syndrome |
| New Referrals | 69.4 | Patient diagnosed with X-linked dominant Kabuki Syndrome 2 due to variant in *KDM6A* was referred to cardiology for an echocardiogram, nephrology for a renal ultrasound, and gastroenterology for feeding difficulties |

\*See full list in Table S5

## DISCUSSION

We describe our experience with genomics-informed neuropsychiatric care from a multidisciplinary specialty clinic for patients with NDDs. Our cohort included a predominance of younger patients, with a peak of toddler- and school-aged youth but including adults into their forties (Fig. S1B). In addition to be housed at a children’s hospital, the enrichment of younger children in our clinic is likely due to the increased rate of genetic testing and diagnosis primarily in pediatric patients, with many older adults with NDDs going untested and undiagnosed. There is a great need to address this disparity through collaboration with adult medical providers so that all patients with NDDs, young and old, can benefit from advances in genomic and precision medicine [18,19]. The majority of patients were assigned male at birth, consistent with the well-established male sex bias for NDDs, especially ASD [20]. Clinic patients tended to have higher SES and white ethnicity, reflective of the region of Los Angeles that the clinic serves, although SES did not correlate with the number of psychiatric diagnoses or having access to next-generation sequencing, suggesting some degree of consistency in diagnostic care independent of SES. Approximately, one-third of patients had comorbid seizure disorders, somewhat higher than reported rates in patients with NDDs [21,22], suggestive of possible referral bias for severely affected individuals or pleiotropic effects of NDD-associated variants.

We observed high rates of baseline psychiatric comorbidities (Table 1), with attention deficit hyperactivity disorder, anxiety disorder, depressive disorder and obsessive compulsive disorder accounting for 83% of psychiatric co-morbidities, consistent with previous reports [23]. The CARING Clinic neurologists, both with behavioral neurology training, initially managed many of these disorders alongside neurological indications such as epilepsy and developmental delays, which is reflected in the high number of neurology visits (Fig. 1). When psychiatric symptoms became extremely acute or treatment refractory or the patient had been referred to The Clinic specifically for psychiatric management, they would be evaluated by psychiatry. This model limited the number of psychiatric visits to serve the largest number of patients. Severe behavioral problems in patients with complex NDDs, including aggression and/or self-harm, can remain acute for extended periods of time requiring high frequency follow-up [24], which we did not have the capacity for as a monthly subspeciality clinic. We attempted to limit psychiatric visits to second-opinion consultations and required all patients to seek care in the community. However, due to the limitation of available psychiatric providers in the community, we would often extend psychiatric care beyond consultation, which further limited CARING Clinic psychiatry access. Insurance denials of psychiatric services, despite authorizing neurological and genetics evaluations for the same patient, proved to be another challenge for providing full multidisciplinary care to patients, consistent with ongoing shortcomings of the Mental Health Parity Law [25].

An alternative clinic model, such as a “hub-and-spoke” model like ECHO Autism [26], where neurogenetic specialists consult with community primary care providers and/or psychiatrists, may allow for more broadly accessible genomics-informed psychiatric care, but with less direct expert engagement. A psychiatry-based clinic with embedded genetic counselors might also be able to provide a higher volume of genomics-informed psychiatric care, although few genetic counselors have extensive training or expertise in psychiatric genetics [27]. Regardless of the model of care delivery, a genomics-informed approach to neuropsychiatric care of patients with NDDs will become increasingly relevant as more patients receive genetic diagnoses and gene-modifying therapies for NDDs come to fruition [28].

We recommended psychoactive medication adjustments for over 60% of patients who were evaluated in CARING Clinic. This may reflect referral bias, as referred patients may have more acute symptoms, the potential treatment-refractory nature of the patient population, and/or the genetics expertise of neuropsychiatric providers with reciprocal collaboration with medical genetics. Many families reported reduced symptoms and improved functioning for complex neuropsychiatric disorders including improved stabilization of schizoaffective disorder in a patient with 22q11.2 deletion syndrome and resolution of treatment-refractory insomnia through suvorexant and quetiapine trials in a patient with Kleefstra Syndrome [29]. No pre- and post-standardized assessment of psychiatric symptoms or adaptive functioning were obtained, limiting our ability to objectively determine the clinical impact of medication adjustments using a genomics-informed approaches compared to treatment-as-usual. This is a future goal of ours.

Chromosomal microarray and Fragile X testing were the most common genetic tests received prior to clinic, consistent with historic professional guidelines [30]. We frequently helped facilitate exome sequencing and chromosomal microarrays (Table 2), resulting in an overall clinic P/LP diagnostic yield of 40.8%. While a few genetic diagnoses were highly prevalent (Table 2) due to dedicated clinics for those conditions within the health system, most patients with an identified P/LP variant had a non-recurrent, ultra-rare genetic disorder (Table S5). Similar observations were recently made in a cohort of 802 children with medical complexity, where 211 out of 265 diagnosed genetic conditions were observed in a single individual and only nine conditions were present in five or more patients [31]. This suggests that genetics education in medical training should consider focusing less on “classic syndromes” such as Down’s Syndrome or Fragile X Syndrome, and instead focus on broad principles of genomic medicine and how to properly leverage genetic resources, such as OMIM [32], Unique [33], and From Genome-to-Treatment [34]. Health systems should also carefully consider the tradeoffs of creating clinics focused on individual neurogenetic disorders versus neurogenetic conditions broadly defined to improve access to specialty care as the number of known, ultra-rare neurogenetic disorders grow. Importantly, the basic needs and clinical presentations of these various rare disorders overlap considerably. By addressing these needs holistically, this could greatly shorten the “diagnostic and therapeutic odyssey” that so many families of youth with NDDs face in trying to get appropriate medical care [12].

Lastly, we demonstrate that taking a genomics-informed approach to neuropsychiatric care for patients with NDDs can affect medical management across multiple domains for most patients (Tables 3 and S5). At very least, additional referrals occur due to known complications associated with a given genetic disorder, but many of our patients also received medication changes secondary to their genetic diagnosis or were able to enroll in clinical trials that they were not previously eligible for. With the rapid expansion of gene-based therapies, making an accurate genetic diagnosis will become ever more critical for our patients to receive proper management and treatment.

## CONCLUSIONS

Our experience suggests that genomics-informed neuropsychiatric care is feasible but complex. We plan to continue optimizing and expanding the multidisciplinary care clinic model at Children’s Hospital Los Angeles to reach a largely underserved population. There are many different potential approaches to genomics-informed neuropsychiatric care, each with potential strengths and limitations. Genetic diagnosis leads to direct changes in clinical management in many cases. Genomics-informed education and care for NDDs should be inclusive and adaptable, as the number of identifiable neurogenetic conditions continues to grow and precision treatments for many neurogenetic disorders are under development.

## Data Availability

The datasets generated and/or analyzed during the current study are not publicly available due to patient privacy laws but are available from the corresponding author on reasonable request.

## LIST OF ABBREVIATIONS

ADI: area deprivation index
ASD: autism spectrum disorders
CARING: care and research in neurogenetics
GDD: global developmental delay
ID: intellectual disability
IRB: institutional review board
NDD: neurodevelopmental disorder
P/LP: pathogenic or likely pathogenic
PNES: psychogenic nonepileptic seizures
SES: socioeconomic status
SVI: social vulnerability index
UCLA: University of California Los Angeles

## DECLARATIONS

### Ethics approval and consent to participate

Research approval was granted by the UCLA Medical Institutional Review Board 3. Legal guardians provided informed consent for prospective collection of clinical data (UCLA IRB#: 14-001908). Patient assent was obtained when feasible. With an IRB-approved waiver of consent, the charts of an additional 206 patients were retrospectively reviewed (UCLA IRB#: 19-000121).

### Consent for publication

Not Applicable

### Authors’ contributions

A.D.B., J.A.M-A. and S.S.J. conceived and designed the study. A.D.B., B.N.S., C.D., R.B.W., S.M., J.A.M.A., and S.J.S. gathered data. D.J.A. and N.R.W. entered and analyzed data. A.D.B. wrote the manuscript. All authors provided critical revision of the manuscript. The authors read and approved the final manuscript.

## Acknowledgements

The authors would like to thank the CARING patients and their families for participating in this research study.

## Authors’ information

Twitter handle: @besterman_a (Aaron Besterman)

