## Supplemental Tables for "Genomics-informed Neuropsychiatric Care for Neurodevelopmental Disorders: Results from A Multidisciplinary Clinic"

**Table S1** Psychoactive Medication Exposures

| CNS Active Medication Class | Ever Exposed | % | Prescribed at Intake | % |
|---|---|---|---|---|
| AED/Mood Stabilizer* | 102 | 32.28 | 69 | 21.84 |
| Alpha Agonist* | 65 | 20.57 | 43 | 13.61 |
| Anticholinergic/Dopamine Agonist | 6 | 1.90 | 5 | 1.58 |
| Atypical Antidepressant* | 16 | 5.06 | 9 | 2.85 |
| Atypical Antipsychotic* | 77 | 24.37 | 52 | 16.46 |
| Barbiturate | 11 | 3.48 | 1 | 0.32 |
| Benzodiazepine* | 61 | 19.30 | 47 | 14.87 |
| Beta Blocker | 5 | 1.58 | 5 | 1.58 |
| First Generation Antihistamine | 13 | 4.11 | 8 | 2.53 |
| First Generation Antipsychotic* | 15 | 4.75 | 9 | 2.85 |
| Lithium | 5 | 1.58 | 1 | 0.32 |
| Melatonin | 47 | 14.87 | 41 | 12.97 |
| Muscle Relaxer/Nerve Pain | 3 | 0.95 | 1 | 0.32 |
| NET Inhibitor | 12 | 3.80 | 6 | 1.90 |
| NMDA Antagonist | 4 | 1.27 | 1 | 0.32 |
| Non-Benzodiazepine Anxiolytic | 2 | 0.63 | 1 | 0.32 |
| Nootropic | 1 | 0.32 | 1 | 0.32 |
| Opioid* | 1 | 0.32 | 1 | 0.32 |
| Opioid Antagonist | 2 | 0.63 | 2 | 0.63 |
| Sedative/Hypnotic* | 2 | 0.63 | 1 | 0.32 |
| SNRI | 4 | 1.27 | 2 | 0.63 |
| SSRI* | 71 | 22.47 | 46 | 14.56 |
| Stimulant* | 57 | 18.04 | 23 | 7.28 |
| TCA | 3 | 0.95 | 2 | 0.63 |
| Triptan | 1 | 0.32 | 1 | 0.32 |

Summary of patient exposures to each of the 25 CNS active medication classes. “Ever” refers to any time prior to date of intake.
*Refers to classes where patients had tried or were taking multiple medications within the same class (i.e., cases of intraclass polypharmacy).

**Table S2** Referrals to Specialized Neurodevelopmental Services from CARING

| | N | % |
|---|---|---|
| **Core Medical Specialties** | | |
| Outside Psychiatry | 10 | 3.16 |
| Outside Neurology | 8 | 2.53 |
| UCLA Autism Evaluation Clinic | 8 | 2.53 |
| Outside Genetics | 6 | 1.90 |
| Developmental-Behavioral Pediatrics | 4 | 1.27 |
| **Specialty Psychiatric and Psychological Services** | | |
| Social Skills Therapy Based Programs | 30 | 9.49 |
| Psychiatric Day Programs | 21 | 6.65 |
| Outside Clinical Psychology and Psychotherapy | 16 | 5.06 |
| Family Therapy, Resources, and Support | 6 | 1.90 |
| Educational and Resource Consultants | 4 | 1.27 |
| Cognitive and Neuropsychological Testing | 2 | 0.63 |
| thinkSMART (executive function training) | 2 | 0.63 |
| Electroconvulsive Therapy | 1 | 0.32 |
| Feeding Therapy | 1 | 0.32 |
| **Condition Specific Clinics and Support Groups** | | |
| Condition Specific Clinics at Outside Institutions | 2 | 0.63 |
| UCLA 22q11.2 Deletion Clinic | 2 | 0.63 |
| Dup15q Alliance | 1 | 0.32 |
| UCLA Cerebral Palsy Clinic | 1 | 0.32 |
| UCLA Child OCD, Anxiety, and Tic Disorders Clinic | 1 | 0.32 |
| UCLA Family Stress, Trauma, and Resilience Clinic | 1 | 0.32 |
| UCLA Muscular Dystrophy Clinic | 1 | 0.32 |
| UCLA Pediatric Acute-onset Neuropsychiatric Syndrome Clinic | 1 | 0.32 |

Number of referrals to medical specialties, clinics, and services specific to the treatment of neurogenetic disorders. Referrals are listed in descending order by frequency within each subcategory.

**Table S3** Referrals to Primary and Subspecialty Medical Care.

| | N | % |
|---|---|---|
| **Primary Care** | | |
| Internal Medicine-Pediatrics | 7 | 2.22 |
| Adolescent Medicine | 2 | 0.63 |
| Internal Medicine | 1 | 0.32 |
| **Medical Subspecialties** | | |
| Gastroenterology | 18 | 5.70 |
| Cardiology/Echocardiography | 15 | 4.75 |
| Nephrology | 6 | 1.90 |
| Pulmonology/Sleep Medicine | 6 | 1.90 |
| Clinical Nutrition/Registered Dietician | 5 | 1.58 |
| Allergy and Immunology | 4 | 1.27 |
| Endocrinology | 4 | 1.27 |
| Rheumatology | 2 | 0.63 |
| Dermatology | 1 | 0.32 |
| East-West (Integrative) Medicine | 1 | 0.32 |
| Hematology/Oncology | 1 | 0.32 |
| **Surgical Subspecialties** | | |
| Orthopedic Surgery and Podiatry | 22 | 6.96 |
| Ophthalmology | 14 | 4.43 |
| Otolaryngology | 4 | 1.27 |
| Urology | 4 | 1.27 |
| Gynecology | 1 | 0.32 |
| Neurosurgery | 1 | 0.32 |
| ***Screenings*** | | |
| Audiology | 8 | 2.53 |
| Optometry | 1 | 0.32 |
| **Other Specialized Medical Testing** | | |
| Renal Ultrasonography | 4 | 1.27 |
| Bone Age Study | 1 | 0.32 |
| DEXA Scan | 1 | 0.32 |
| Swallow Study | 1 | 0.32 |

Number of referrals to primary care and to subspecialties required to manage medical comorbidities of neurogenetic disorders. Referrals are listed in descending order by frequency within each subcategory.

### **Table S4** Genetic Diagnoses Identified in Individual Patients

| Primary Genetic Diagnoses | VUS |
|---|---|
| 10p22 and 10q23 Deletion Syndrome | |
| 14q11.2 Duplication Syndrome | |
| 15q13.3 Microdeletion Syndrome | |
| 15q15.2q21.1 Deletion Syndrome | |
| 16p13.11 Microduplication Syndrome | |
| 16p13.22p12.3 Duplication Syndrome | |
| 16q11.2 Deletion Syndrome | |
| 17p12 Deletion Syndrome (includes *PMP22*) | |
| 18p Deletion Syndrome | |
| 18q21.3 Deletion Syndrome | |
| 19q13.12 Microdeletion + 19p13 Duplication (including *PRKACA*) Syndromes | |
| 1p36.12-13 Duplication Syndrome | 22q11.22 duplication |
| 22q11.23 Duplication Syndrome | *CNGA2*, *PLXNA3*, and *MTHFR* |
| 2q22.2q23.3 Deletion Syndrome | |
| 3p21.31 Microdeletion Syndrome | |
| 3q29 Duplication Syndrome | |
| 6q26q27 Inverted Triplication Syndrome | |
| 7q11.2 Deletion (including *AUTS2*) Syndrome | |
| 7q35 Deletion Syndrome | |
| 8p23.3p23.1 Deletion and 8p23.1p11.21 Duplication | |
| 9p22.3 Microdeletion Syndrome and Gorlin Syndrome | |
| 9q21.13 Deletion Syndrome (includes *RORB*) | |
| 9q34.11 Deletion Syndrome | |
| Autosomal Dominant Familial Hypercholesterolemia | |
| Autosomal Dominant Genitopatellar Syndrome (*KAT6B*) + Autosomal Dominant 46XY Disorder of Sexual Development (*MAP3K1*) | |
| Autosomal Dominant Intellectual Disability (*FOXP1*) | |
| Autosomal Dominant Intellectual Disability (*PURA*) | |
| Autosomal Dominant Intellectual Disability 23 | |
| Autosomal Recessive Deafness (*GJB3* Gene H91.8X9 Mutation) | |
| Bannayan-Riley-Ruvalcaba Syndrome | |
| Bohring-Opitz Syndrome | |

| **Primary Genetic Diagnoses** | **VUS** |
|---|---|
| Brachydactyly Intellectual Disability Syndrome | |
| *BRWD3* | *CHD8,* 3p26.3 and 10p15.315.2 duplications, X-skewing |
| *CACNA1C* | Xp22.31 (including *WWOX*) deletion |
| *CDKL* | |
| *CHD2* | *HDAC8* |
| Chromosome 1 Deletion Syndrome | |
| *CLTC* | 1p31.1 and 5q21.3 duplications |
| Coffin-Lowry Syndrome | |
| Cornelia De Lange Syndrome | |
| Cowden Syndrome | |
| *DDX3X* variant | |
| *DEAF1* variant | |
| *DHX30* variant | |
| Down Syndrome | |
| Dravet Syndrome | |
| Duchenne Muscular Dystrophy | |
| Duplication 16p11.2 Syndrome | 7q31.33 deletion |
| Factor V Leiden | MTHFR |
| *FGF3* | 8q23.1 duplication |
| *FOXG1* | |
| Galactosemia + 4q12 deletion + 17q11.2 duplication | |
| *GATAD2B* deletion + *TBC1D7* | 8p23.2 deletion |
| *GRIN1* variant | |
| Helsmoortel-Van Der Aa Syndrome | |
| Hunter Syndrome | |
| Hypomyelinating Leukodystrophy Type 12 | |
| *KIF11* variant with additional variants of uncertain significance in | *CDCA5* and *MT-ND5* with 8% heteroplasmy |
| Klinefelter Syndrome | |
| Lowe Syndrome | |
| Monosomy 9p2 | |
| Mosaic Supernumerary 9p13.3q13 Duplication Syndrome | |
| Mosaic Turner Syndrome | |
| Noonan Syndrome | |
| *NRAS* variant | |
| *NRXN1* Deletion Syndrome | |
| Opitz G/BBB Syndrome | |
| Partial Monosomy 2q | |

| Primary Genetic Diagnoses | VUS |
|---|---|
| Partial Trisomy 18q | |
| Phelan-McDermid Syndrome | |
| *POLR3B* variant with additional *SYNE1* variant and gray zone Fragile X repeats (49) | |
| Potocki-Lupski Syndrome | |
| Rett Syndrome | |
| Say-Barber-Biesecker-Young-Simpson Syndrome with Ehlers-Danlos Syndrome Type 1 | |
| *SET5D* variant with additional *KIF1A* variant of uncertain significance | |
| Smith-Lemli-Opitz Syndrome | |
| Smith-Magenis Syndrome | |
| Snyder-Robinson Syndrome | |
| Stickler Syndrome | |
| Triple X Syndrome | |
| *TTN* variant with additional *KIF7* variant of uncertain clinical significance | |
| Ulrich Myopathy (*COL6A1*) | |
| Unbalanced t(8;12) | |
| VACTERL Association | |
| Xp11.23P11.22 Microduplication Syndrome | |
| Xp22.33p22.31 Deletion Syndrome | |
| *ZNF462* variant | |

### **Table S5:** Full Management Implications for Patients Diagnosed with Genetic Disorders in CARING Clinic

| Genetic Finding for Patient | Management Implications |
|---|---|
| 3p25 | Referrals for echocardiogram, renal ultrasound, ENT consult and re-evaluation of myotonia by neuromuscular specialist. Ongoing surveillance for seizures. |
| TSC (clinical) | Enrolled in diagnostic clinical study to search for intronic TSC variants |
| *TTN*, *KIF7* | Patient and father (whom both carried variant) receive echocardiograms. Ophthalmology referral to assess for optic atrophy and retinal pigmentation. Referral to diagnostic clinical study. |
| *ADNP* | Referred for sleep study, ophthalmology, cardiology, nephrology, and for brain MRI. Enrolled in ketamine infusion trial and referred to gene therapy trial as well. Psychotropic medications selected to avoid exacerbating potential known medical complications. |
| *KDM6A* | Referred for echocardiogram, renal ultrasound, and gastroenterologist. Had a re-evaluation of hypotonia by clinic neurologists. |
| *MECP2*, *HMBS* | Referrals to developmental pediatrics, gastroenterology, orthopedics for scoliosis evaluation, sleep apnea evaluation, EKG to assess QTc, vEEG to help decide if anti-epileptics were necessary. Reproductive counseling provided. |
| *BRD4* | Referrals to developmental pediatrics for autism evaluation, referral to cardiology for echocardiogram and assessment of thrombocytopenia, referred to social work to discuss long-term care. Referred to online family group. |

| | |
|---|---|
| *SUMF1* | Referred to lysosomal storage disease clinic. Recommended echocardiogram, sleep and swallow studies, abdominal ultrasound. Referred to nutritionist at metabolic clinic, gastroenterologist and, and ophthalmology. |
| *SUMF1* | Referred to lysosomal storage disease clinic. Recommended echocardiogram, sleep and swallow studies, abdominal ultrasound. Referred to nutritionist at metabolic clinic, gastroenterologist and, and ophthalmology. |
| *SUMF1* | Referred to lysosomal storage disease clinic. Recommended echocardiogram, sleep and swallow studies, abdominal ultrasound. Referred to nutritionist at metabolic clinic, gastroenterologist and, and ophthalmology. |
| *GATAD2B* | Enrolled in diagnostic clinical stud, reproductive counseling, referred for autism evaluation |
| 9q34.11 | Referred for brain MRI, EEG, and further evaluation of SMA at neuromuscular clinic. |
| 8q23.1, *FGF3* | Referred for abdominal and brain MRIs, dental clinic, social work for programs for patients who are hard of hearing. Patient referred to schannomatosis clinic at for surveillance. Recommended paternal family members and siblings undergo surveillance for early-onset cancers. |
| *FOXP1* | Referred to gastroenterology, neurology, developmental pediatrics, cardiology, and nephrology. |
| *CHD2*, *HDAC8* | Referred to child psychology for cognitive testing and the SIMONS VIP study and UCLA clinical research study. Increased surveillance with EEG for seizures. |
| *DHX30* | Referral to neuromuscular clinic, gastroenterology, orthopedics, and physical and occupational therapy. Obtained repeat brain MRI. |
| *PTEN* | Referred for thyroid ultrasound to monitor for cancer. Referred for behavioral therapies including ABA. Referred to neurology and dermatology for initiation of Everolimus. |
| Opitz G/BBB syndrome | Referred to ENT for management of laryngeal cleft. Patient ultimately received surgery that helped with breathing symptoms. Referred to gastroenterology, cardiology and ophthalmology. |
| 2q22.3 q23.3 | Referred for hand x-rays to assess for anomalies seen in potential Coffin-siris syndrome, including hypoplastic middle and distal phalanges. Recommended testing of grandparents. Able to explain that transient cardiomyopathy and recurrent ear infections and UTIs were related to genetic disorder (but did not initiate new referrals). Recommended family member get evaluated for further phenotypic characterization. |
| *SMS* | Referral to Undiagnosed Diseases Program, gastroenterology, nephrology, and orthopedics. Referral for neurodevelopmental evaluation and behavioral therapy, and recommended parents connect with other families on social media for community support. |
| *COL6A1* | Referral to neuromuscular specialist |
| 15q11.2q13.3 | Referral to clinical research studies. Encouraged to participate in dup15q family conference, connected to families with same condition, aggressive monitoring for development of seizures, which guided recommendations for anti-epileptic medication management. |
| 8p23.3p23.1, 8p23.1p11.21 | Referral to ophthalmology, developmental pediatric, cardiology, nephrology, brain MRI, orthopedic surgery, and a UCLA clinical research study. |
| *ZNF462* | Based on GeneMatcher findings, recommended surveillance for leukemia and make referral to endocrinology. Recommended echocardiogram, abdominal ultrasound, renal ultrasound, regional center connection and services, intensive behavioral services, feeding therapy |
| 18q21.3 | Mother tested and given reproductive counseling. Referral to urology, orthopedics, cardiology, nephrology, ophthalmology, cerebral palsy clinic, regional center and early intervention. |
| 9p13.3q13 | Provided reproductive counseling to family members. Encouraged mother to seek out community support on social media. |
| *CLTC*, 1p31.1, 5q21.3 | Reproductive counseling. Referred to diagnostic clinical research study. |
| *SETD5*, *KIF1A* | Genetic counseling. |
| Xp22.33>Xp22.31 | Reproductive counseling for parents. Close surveillance of seizure onset. |
| *LDLR* | Referred to cardiology, ataxia clinic, and clinical research study. |

| | |
|---|---|
| *GNAQ*, *PIK3CA* | Referral to overgrowth syndrome clinic for dental and orthopedic care. |
| *RAI1* (17p11.2), *MYO15A* | Brain MRI, clarified that patient is not at risk of neuropathy, advised prenatal testing should be discussed with obstetrician, speech evaluation. |
| *NRAS* | EKG, echocardiogram, CBC, TSH, repeat cardiac exams in adolescence, monitor for symptoms of lymphatic complications and seizures. |
| *PURA* | Referral to cerebral palsy clinic and sleep study. Surveillance for seizures. Consideration of dopaminergic medications, and referral for intensive behavioral services for developmental delays and hypotonia. |
| *POLR3B* and *SYNE1* | Parental genetic testing. Evaluation for Tourette's that may be genetically linked. Creatine kinase level through primary care doctor. |
| *F5* | Shared results with family and provided education, documentation limited on further impact on management. |
