## Supplemental Figures for "Genomics-informed Neuropsychiatric Care for Neurodevelopmental Disorders: Results from A Multidisciplinary Clinic"

**Figure S1** Patient Demographics

(A)

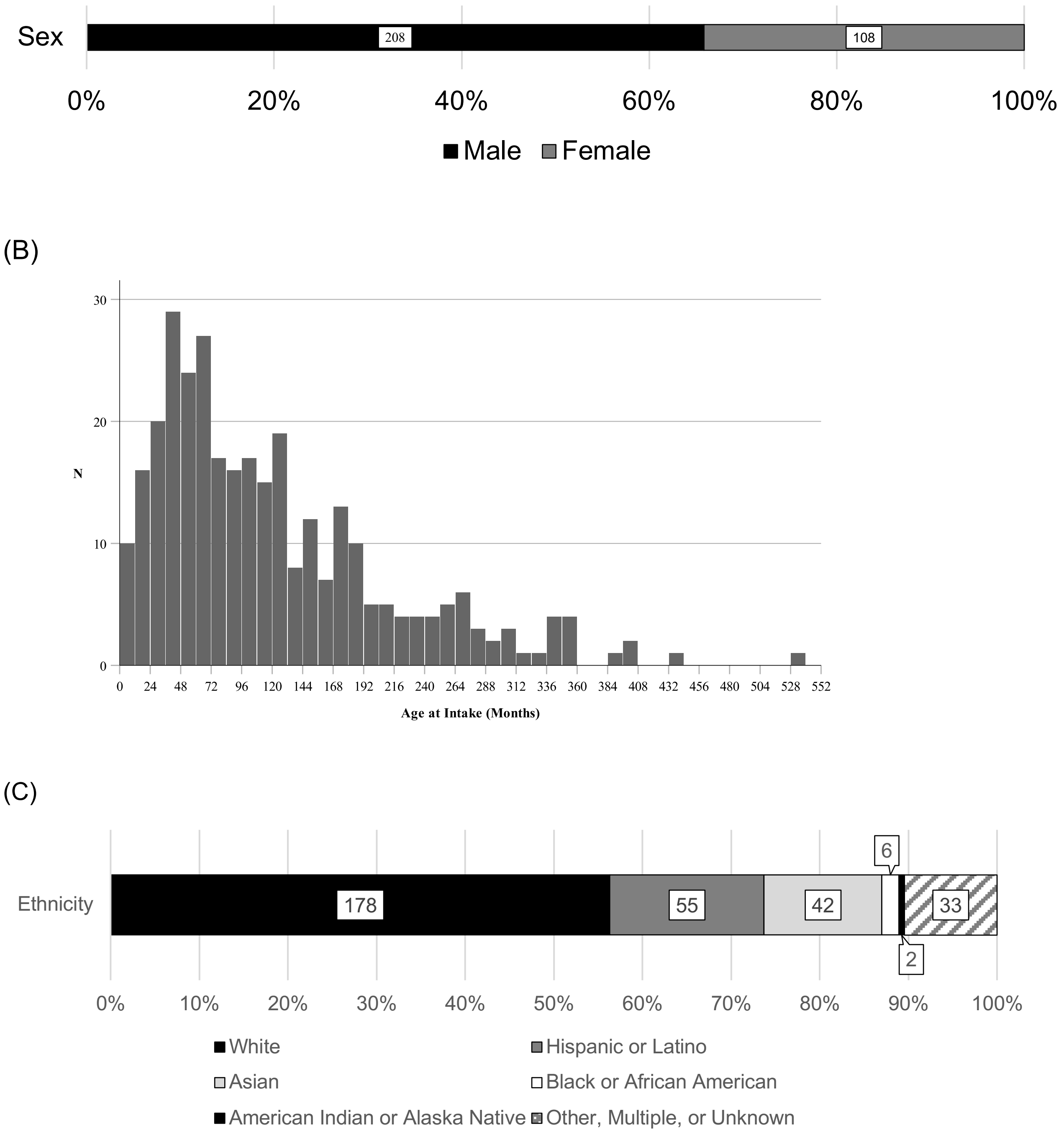


(A) Distribution of patient assigned sex at birth. (B) Histogram showing the age distribution of our patient cohort. Each bin represents a 12-month period ($M$ = 118.99 months, $SD$ = 92.69, range 0-535). (C) Distribution of patient ethnicity by NIH category.

**Figure S2**

(A)

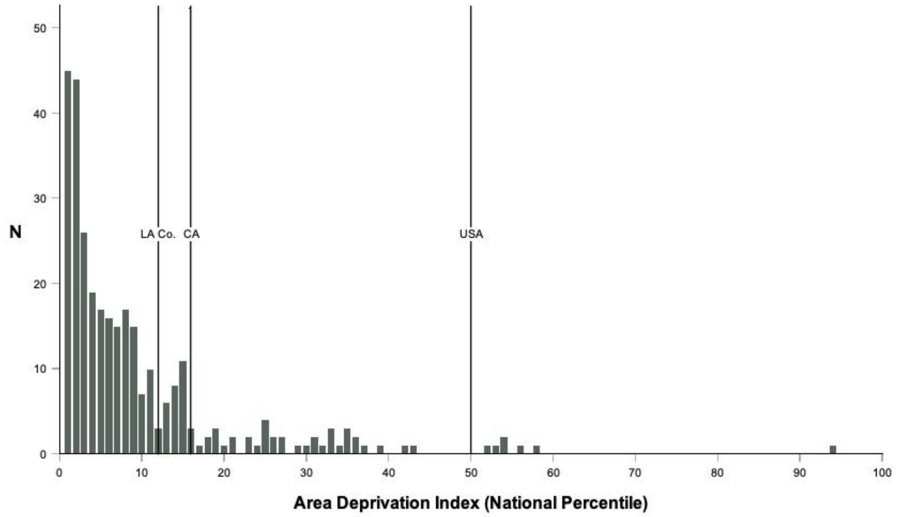


(B)

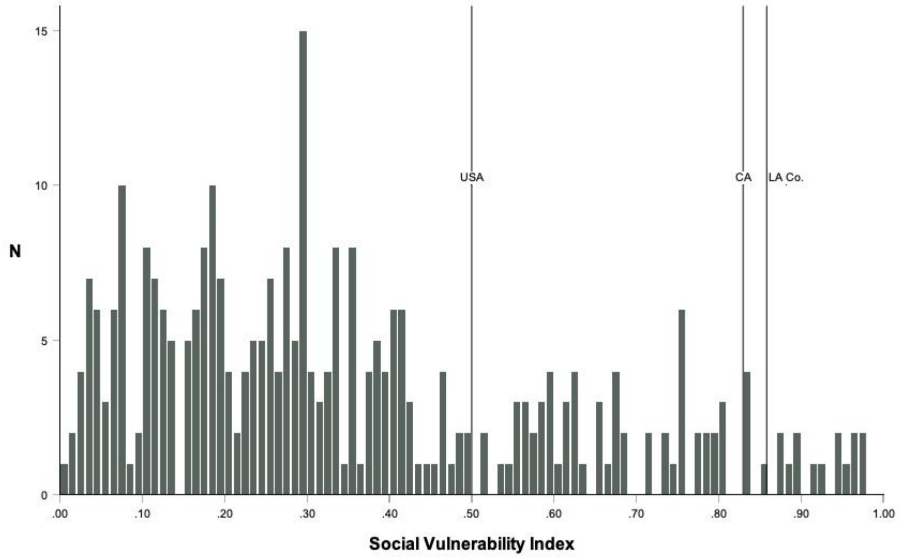


(A) Area deprivation Index (ADI) and (B) Social Vulnerability Index (SVI) are two measures of socioeconomic status (SES). ADI and SVI are inversely correlated with SES. Based on their reported address, most CARING patients fell well below the national (USA), California state (CA), and Los Angeles County (LA Co.) mean for both ADI and SVI.

**Figure S3** Psychoactive Medication Prescription History

(A)

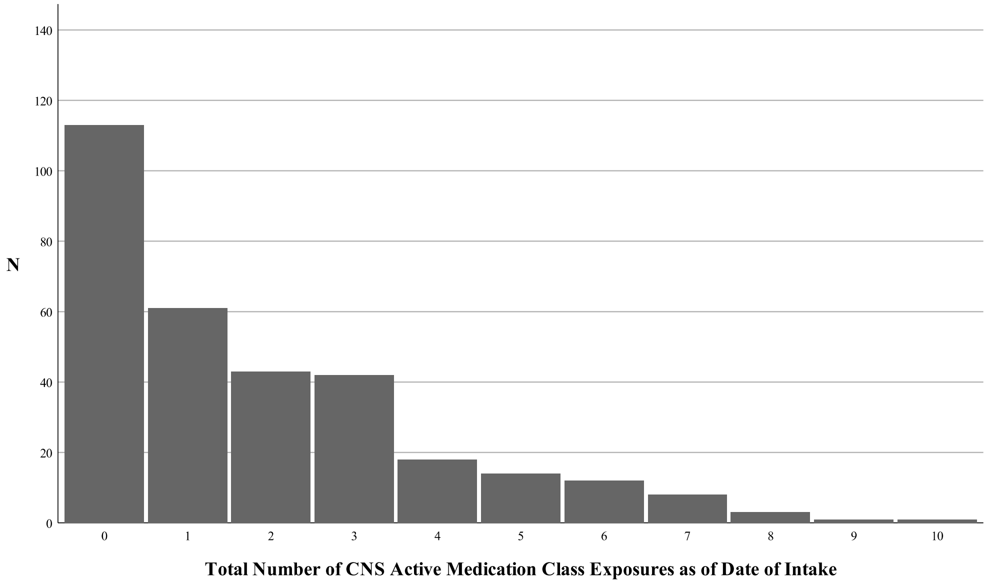


(B)

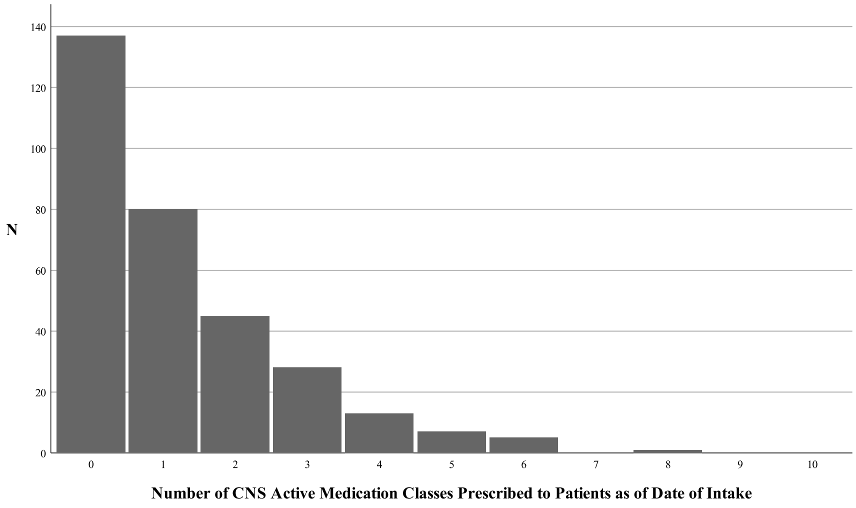


(A) Total number of CNS active medication class exposures as of the date of intake. Total represents the sum of unique classes trialed (but no longer taking on date of intake) and unique classes prescribed as of the date of intake. (B) Total number of CNS active medication class exposures prescribed to the patient by any provider as of the date of intake.

**Figure S4** Other Medical and Surgical Comorbidities.

(A) Without Surgical History

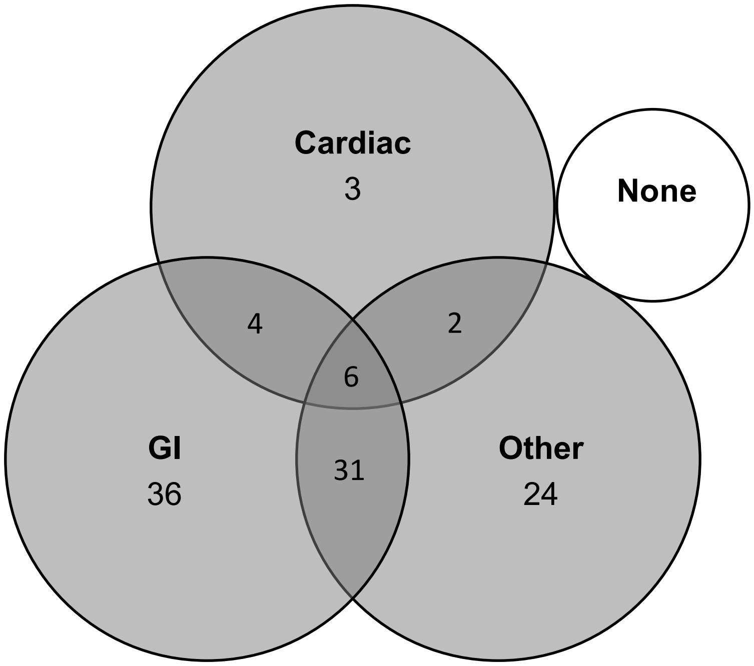


(B) With Surgical History

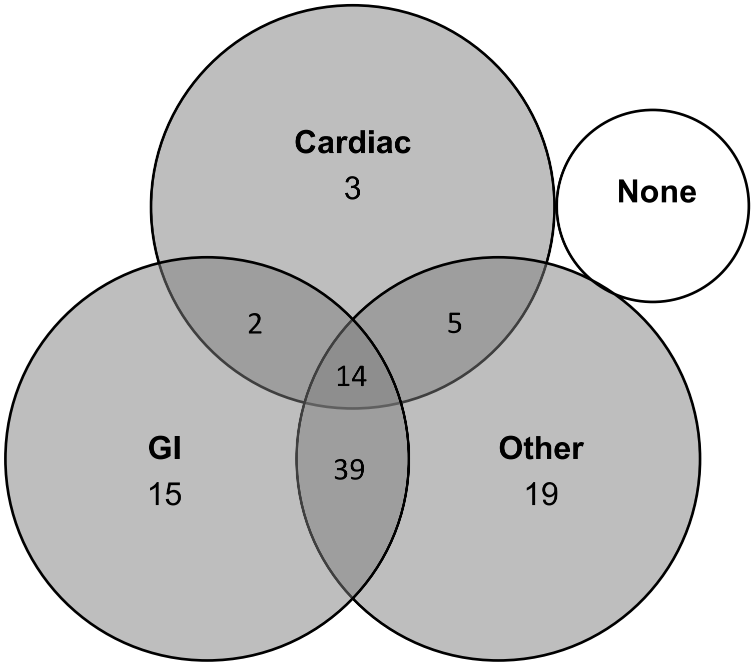

Sum of all numbers within each circle provides the total number of patients with each comorbidity. The numbers within regions of overlap represent the number of patients with combinations of multiple types of comorbidities. GI morbidities include GERD, constipation, diarrhea, and other. Cardiac morbidities include structural cardiac disease and isolated heart murmurs. Other medical comorbidity includes any non-neuropsychiatric, non-GI, non-cardiac condition. Groups represented in panels (A) and (B) are divided by surgical history, which includes any documented surgeries, regardless of complexity.

**Figure S5** Summary of Service Usage

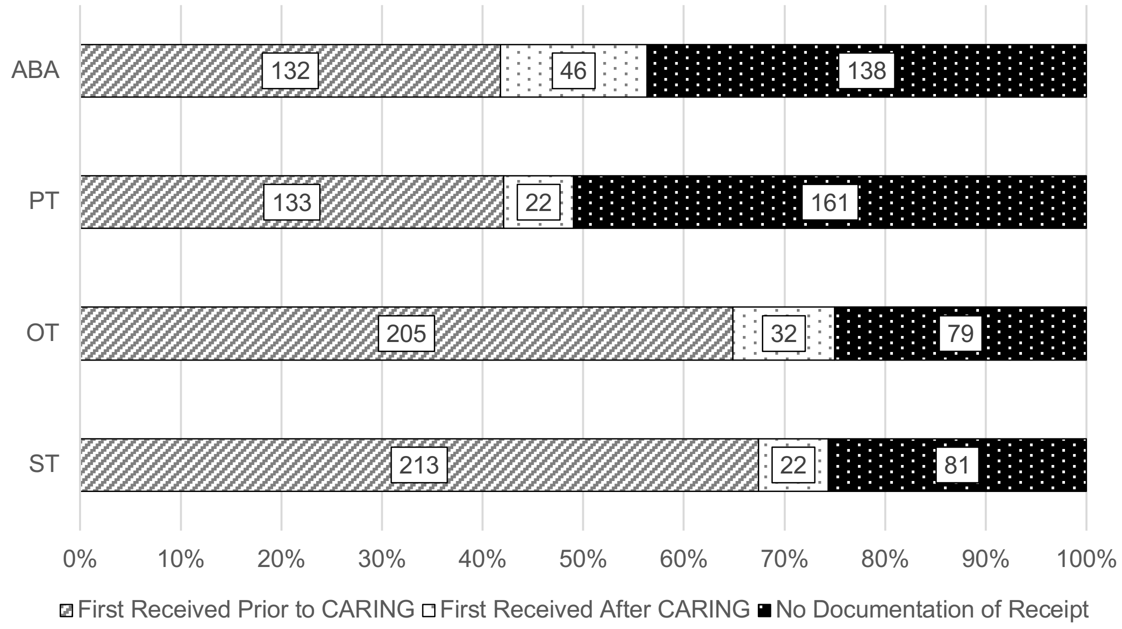


ABA = Applied Behavioral Analysis; OT = Occupational Therapy; PT = Physical Therapy; ST = Speech Therapy
